# A human-interpretable machine learning approach to predict mortality in severe mental illness

**DOI:** 10.1101/2021.04.05.21254684

**Authors:** Soumya Banerjee, Pietro Liò, Peter B. Jones, Rudolf N. Cardinal

## Abstract

Machine learning (ML), one aspect of artificial intelligence (AI), involves computer algorithms that train themselves. They have been widely applied in the healthcare domain. However, many trained ML algorithms operate as “black boxes”, producing a prediction from input data without a clear explanation of their workings. Non-transparent predictions are of limited utility in many clinical domains, where decisions must be justifiable.

Here, we apply class-contrastive counterfactual reasoning to ML to demonstrate how specific changes in inputs lead to different predictions of mortality in people with severe mental illness (SMI), a major public health challenge. We produce predictions accompanied by visual and textual explanations as to how the prediction would have differed given specific changes to the input. We apply it to routinely collected data from a mental health secondary care provider in patients with schizophrenia. Using a data structuring framework informed by clinical knowledge, we captured information on physical health, mental health, and social predisposing factors. We then trained an ML algorithm and other statistical learning techniques to predict the risk of death.

The ML algorithm predicted mortality with an area under receiver operating characteristic curve (AUROC) of 0.80 (95% confidence intervals [0.78, 0.82]). We used class-contrastive analysis to produce explanations for the model predictions. We outline the scenarios in which class-contrastive analysis is likely to be successful in producing explanations for model predictions. Our aim is not to advocate for a particular model but show an application of the class-contrastive analysis technique to electronic healthcare record data for a disease of public health significance.

In patients with schizophrenia, our work suggests that use or prescription of medications like antide-pressants was associated with lower risk of death. Abuse of alcohol/drugs and a diagnosis of delirium were associated with higher risk of death. Our ML models highlight the role of co-morbidities in determining mortality in patients with SMI and the need to manage them. We hope that some of these bio-social factors can be targeted therapeutically by either patient-level or service-level interventions. Our approach combines clinical knowledge, health data, and statistical learning, to make predictions interpretable to clinicians using class-contrastive reasoning. This is a step towards interpretable AI in the management of patients with SMI and potentially other diseases.

## Introduction

In this article we apply a recent development in machine learning, termed class-contrastive analysis, to the major public health problem of premature mortality in schizophrenia. Schizophrenia affects approximately 0.5% of the population [1]. It is a severe mental illness (SMI), along with bipolar affective disorder, personality disorders, and recurrent depressive disorder. Patients with schizophrenia or other SMI more broadly have substantially increased mortality and reduced life expectancy, often due to physical comorbidities [2] [3] [4] [5]. A challenge for clinical practice is therefore to identify patients at particularly high risk of adverse events (including premature death) and seek to intervene early.

Machine learning (ML) offers a potential route to risk prediction in the field of SMI as elsewhere. ML, a sub-field of artificial intelligence (AI), involves computer algorithms that automatically adjust in response to training data (“train themselves”). Their attractiveness relates to their ability to create predictive models from complex data sets with minimal human intervention, to a degree that may exceed the accuracy of classical statistical models such as logistic regression. ML achieves this through a variety of techniques. In a basic technique such as a multi-layer artificial neural network, for example, a layer of hidden nodes is trained by the algorithm to respond to weighted combinations of inputs; there may be further such layers responding to weighted combinations of the first layer, and these layers may interact recurrently. The “output” layer, giving the prediction or classification, responds to weighted combinations of preceding nodes. The algorithm seeks to minimize output prediction error. As a result, the output may predict accurately (if validated on independent data to avoid overfitting), but it may be very hard for a human to discern how the decision was reached. Clinically, it may be impractical to rely on such a black box predictor.

Here, we develop ML models of mortality in schizophrenia and apply the technique of class-contrastive reasoning to improve their explicability. Class-contrastive reasoning is a technique from the social sciences [7]: the contrast is to an alternative class of exemplars. An example of a class-contrastive explanation is: “The selected patient is at high risk of mortality because the patient has dementia in Alzheimer’s disease and has cardiovascular disease. If the patient did not have both of these characteristics, the predicted risk would be much lower.”

We apply an ML and class-contrastive framework to data on clinically relevant bio-social variables spanning physical health, mental health, personal history, and social predisposing factors. We collate this information from an electronic clinical records system and use clinician knowledge to transform them into features that are used to train an ML system (Fig. 1). We use the ML model to predict mortality and apply class-contrastive reasoning to explain the model.

**Figure 1.**
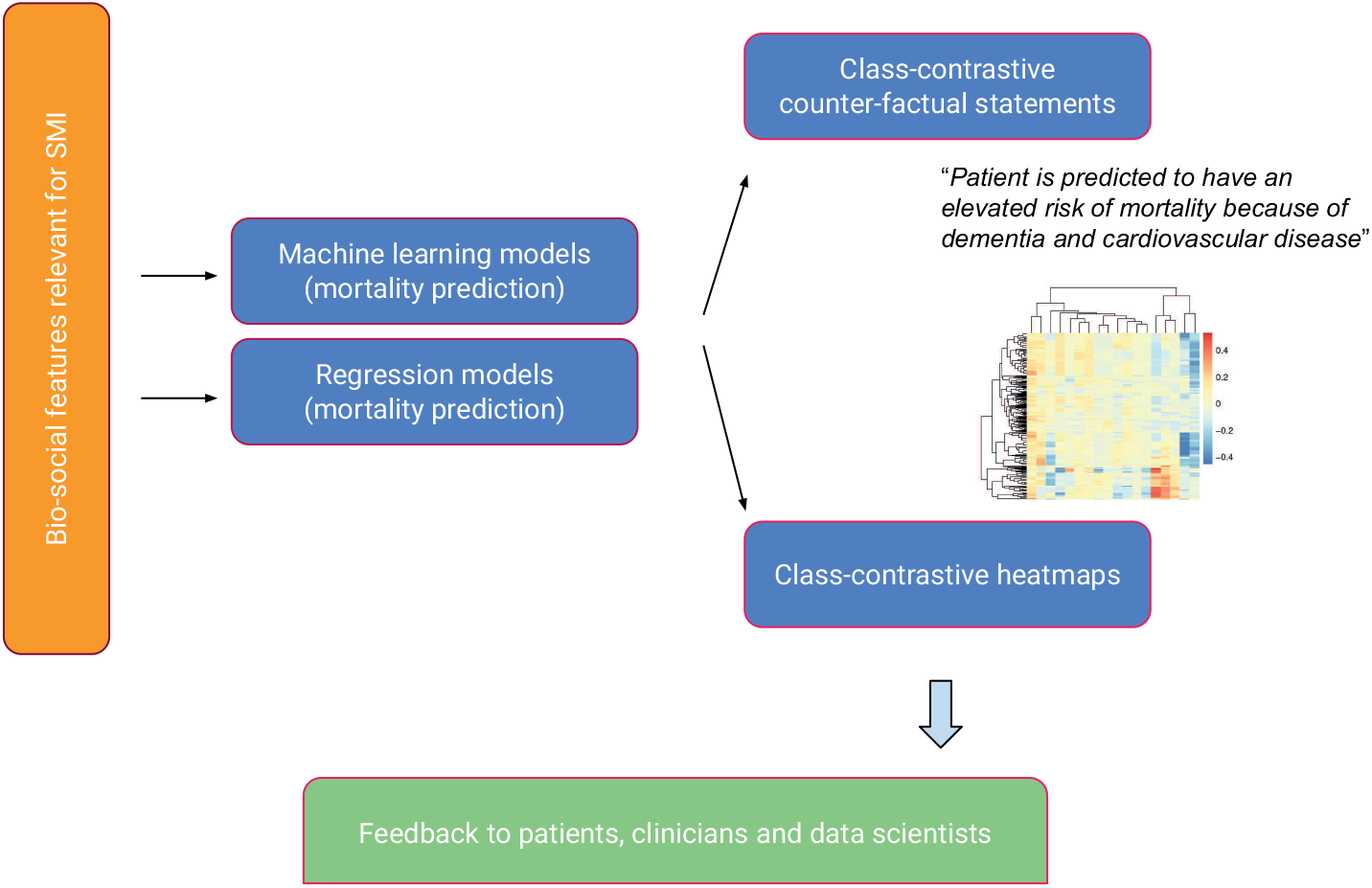
Overview of approach. Bio-social factors that are relevant in severe mental illnesses (SMI) are derived from the electronic healthcare record system and used as inputs to statistical and machine learning algorithms. The algorithms are made interpretable using class-contrastive reasoning. The class-contrastive textual statements and heatmaps aid the understanding of models by domain experts such as clinicians, patients, and data scientists.

We also use a visualization technique for machine learning models (class-contrastive heatmaps) that allows us to map the effect of changing a set of features.

Our approach can be helpful when explicit causal structure is modelled, and when there are a few features which are binary (categorical) in nature. Our approach may not be successful when there are continuous features or many (hundreds) features. If features are hard to define and have to be discovered (for example, by semi-supervised techniques), our approach may not be helpful. Most of our features are binary (categorical) in nature and hence the class-contrastive approach can be applied successfully.

The class-contrastive approach may also be used to evaluate the practical limits of explainability of some models. For example, if a model has hundreds or thousands of features, it may be computationally intractable to exhaustively explore how changing combinations of these features affects the model output.

Our aim is not to advocate for a particular statistical model or black box model. Our objective is to give an example of how class contrastive reasoning can be used to explain black box models with binary categorical features in real world electronic healthcare record data, in a disease of public health significance.

Our aim is not to exhaustively compare all possible statistical models but merely briefly survey and analyse some techniques. We note that our aim is not to demonstrate that some machine learning models can perform better than others.

We show a practical demonstration on a clinical dataset in a disease of public health relevance. We also outline the instances in which class-contrastive reasoning can be successfully applied to electronic health care record data. We suggest class-contrastive reasoning as a method to begin understanding ML and statistical models that have non-linearities. To the best of our knowledge this is the first application of this technique to real world electronic healthcare record data.

Our work is a step towards personalised medicine and interpretable AI in mental health and has the potential to be applicable more broadly in healthcare.

## Data and Methods

### Overview of Methods

We give a brief overview of our approach in this section. Our approach is summarised in Fig. 1.

1. We take de-identified data from an electronic patient record system for mental health.
2. We define a set of high-level features that are in this example time independent. These include age, diagnostic categories (time-independent coded diagnosis at any point during the study period), and medication categories (time-independent prescription of or use of medications). We also include bio-social factors that are important in SMI like information on mental health diagnosis, relevant risk history such as a prior suicide attempt, substance abuse, and social factors such as lack of family support.
3. We use these features to predict death during the time of observation.
4. We use classical statistical models including logistic regression, survival models, and standardised mortality ratios.
5. We then fit machine learning models, comparing predictive accuracy to the classical statistical models.
6. Class-contrastive heatmaps are used to visualize the explanations of the statistical models and machine learning predictions. The corresponding class-contrastive statements also aid human interpretation.

### Mental health clinical record database

We used data from the Cambridgeshire and Peterborough NHS Foundation Trust (CPFT) Research Database. This comprises electronic healthcare records from CPFT, the single provider of secondary care mental health services for Cambridgeshire and Peterborough, UK, an area in which approximately 856,000 people reside. The records are de-identified using the CRATE software [8] under NHS Research Ethics approval (12/EE/0407, 17/EE/0442).

Data included patient demographics, mental health and physical co-morbidity diagnoses: these were derived from coded ICD-10 diagnoses and analysis of free text through natural language processing (NLP) tools [9] [10].

Dates of death are automatically updated via the National Health Service (NHS) Spine. We considered all patients with coded diagnoses of schizophrenia who had records in the electronic healthcare system from 2013 onwards. There were a total of 1706 patients diagnosed with schizophrenia defined by coded ICD-10 diagnosis (diagnosis code F20). We note there is under-coding of schizophrenia.

### Medicine information on prescribed drugs

We extracted medicine information for each patient by using natural language processing on clinical free text data using the GATE software [9] [11].

### Population mortality data

Population mortality data for England and Wales was used from the Office for National Statistics (ONS) [12].

### Data input to statistical algorithms

The features fed in to our statistical and machine learning algorithms included age, gender, high-level diagnosis categories and medication categories. We also included other bio-social factors important in SMI. All these features are used to predict mortality. The full list of features was as follows:

1. High-level medication categories were created based on domain-specific knowledge from a clinician [RNC]. These medication categories are: second-generation antipsychotics (SGA: clozapine, olanzapine, risperidone, quetiapine, aripiprazole, asenapine, amisulpride, iloperidone, lurasidone, paliperidone, sertindole, sulpiride, ziprasidone, zotepine); first-generation antipsychotics (FGA: haloperidol, benperidol, chlorpromazine, flupentixol, fluphenazine, levomepromazine, pericyazine, perphenazine, pimozide, pipotiazine, prochlorperazine, promazine, trifluoperazine, zuclopenthixol); antidepressants (agomelatine, amitriptyline, bupropion, clomipramine, dosulepin, doxepin, duloxetine, imipramine, isocarboxazid, lofepramine, maprotiline, mianserin, mirtazapine, moclobemide, nefazodone, nortriptyline, phenelzine, reboxetine, tranylcypromine, trazodone, trimipramine, tryptophan, sertraline, citalopram, escitalopram, fluoxetine, fluvoxamine, paroxetine, vortioxetine and venlafaxine); diuretics (furosemide); thyroid medication (drug mention of levothyroxine); antimanic drugs (lithium) and medications for dementia (memantine and donepezil).
2. Relevant co-morbidities we included were diabetes (inferred from ICD-10 codes E10, E11, E12, E13 and E14 and any mentions of the drugs metformin and insulin), cardiovascular diseases (inferred from ICD-10 diagnoses codes I10, I11, I26, I82, G45 and drug mentions of atorvastatin, simvastatin and aspirin), respiratory illnesses (J44 and J45) and anti-hypertensives (mentions of the drugs bisoprolol and amlodipine).
3. We included all patients with a coded diagnosis of schizophrenia (F20). For these patients with schizophrenia, we also included any additional coded diagnosis from the following broad diagnostic categories: dementia in Alzheimer’s disease (ICD-10 code starting with F00), delirium (F05), mild cognitive disorder (F06.7), depressive disorders (F32, F33) and personality disorders (F60).
4. We also included relevant social factors: lack of family support (ICD-10 chapter code Z63) and personal risk factors (Z91: a code encompassing allergies other than to drugs and biological substances, medication noncompliance, a history of psychological trauma, and unspecified personal risk factors); alcohol and substance abuse (this was inferred from ICD-10 coded diagnoses of Z86, F10, F12, F17, F19 and references to thiamine which is prescribed for alcohol abuse). Other features included are self-harm (ICD-10 codes T39, T50, X60, X61, X62, X63, X64, X78 and Z91.5), non-compliance and personal risk factors (Z91.1), referral to a crisis team at CPFT (recorded in the electronic healthcare record system) and any prior suicide attempt (in the last 6 months or any time in the past) coded in structured risk assessments.

These broad categories constituted our representation of simplified clinician-based knowledge. We use these features (including age of the patient) to predict whether a patient died any time during the time period observed (from first referral to CPFT to the present day). We do not attempt to predict the risk of dying, for instance, 1 year after first referral to CPFT. The features we use to predict mortality are also time-independent. This represents a simplified time-independent model. More detailed modelling would include temporal effects of such predictors.

Age is a predictor in all our models, including survival models. We consider time of death and time of feature collection. The observed outcome (death) was binary and this is the outcome the models are predicting but models do so via a continuous variable related to risk/probability, so this is simultaneously predicted. Our model predictions, if independently validated in another clinical setting, can be converted in to a risk or probability.

All our models, including the machine learning model, include age as a predictor. However the class-contrastive analysis and the class-contrastive heatmaps do not include age since the feature changes (one at a time or pairwise) can be achieved only for binary (categorical) features. Hence, the class-contrastive heatmaps show the effect of changing predictors on the model prediction, over and above the contribution of age.

### Data pre-processing

Diagnostic codes were based on the International Classification of Diseases (ICD-10) coding system [13]. Age of patients was normalised (feature scaled) by subtracting the mean age from the age of each patient and then dividing by the standard deviation. All categorical variables, such as diagnosis and medications (described above), were converted using a one-hot encoding scheme. This is explained in detail in the Supplementary Section.

### Machine learning and statistical techniques

We performed logistic regression using generalized linear models [14] [15]. We used age (feature scaled) as a continuous predictor. There are categorical features (medications, co-morbidities and other social and personal predisposing factors) which are encoded using a one-hot representation. Additional details are available in the Supplementary Section.

For our machine learning approach, we used artificial neural networks (autoencoders) to integrate data from different sources giving a holistic picture of mental health, physical health and social factors contributing to mortality in SMI. We use the same set of features for all algorithms.

Artificial neural networks are composed of computational nodes (artificial neurons) that are connected to form a network. Each artificial neuron performs a simple computation (much like logistic regression). The neurons are organised in layers. The input layer takes in the input features, transforms them, and passes it to one or more intermediate layers called hidden layers. The hidden layer performs further transformations and passes the result to the output layer. The final output layer is used to make a prediction (in this case, about mortality).

The autoencoder is a type of artificial neural network that also performs dimensionality reduction since the hidden layer has fewer neurons that the input layer [16]. In our framework, the reduced dimensions of the autoencoder (output of the hidden layer) were used as input to a random forest model to predict mortality (Fig. 2). Random forests are machine learning models that build collections of decision trees [17]. Each decision tree makes a prediction after making a series of choices based on the input data. These decision trees are combined to build a collection (forest) that together has better predictive ability than a single tree.

**Figure 2.**
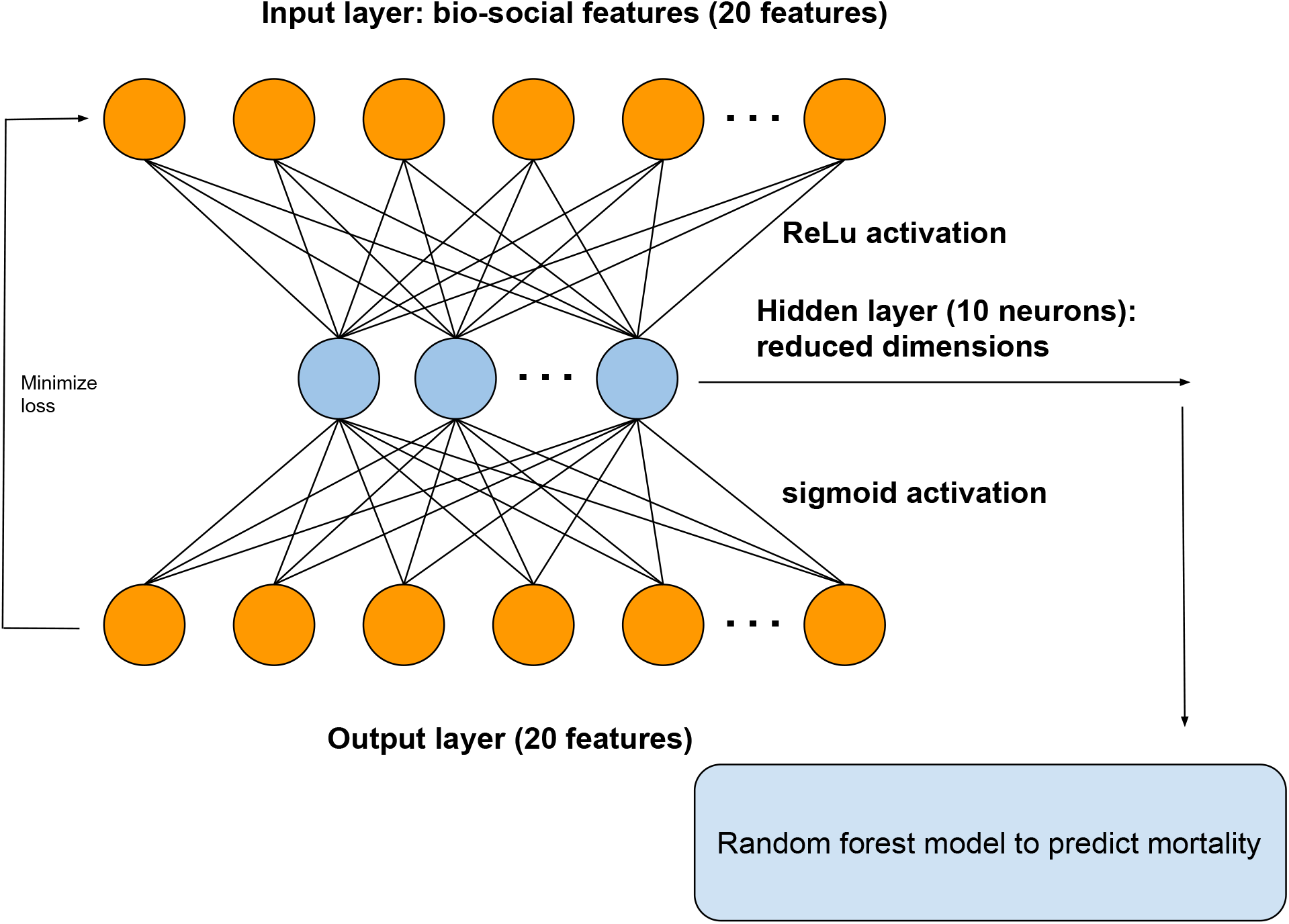
Architecture of autoencoder. The autoencoder takes as input the bio-social features. The output layer is used to reconstruct the input. The hidden layer of the autoencoder is used for dimensionality reduction. We use the hidden layer as input to a random forest model to predict mortality. The hidden layer is composed of 10 neurons.

We split the data into training set (50%), validation set (25%) and test set (25%). We performed 10-fold cross-validation and regularization to penalize for model complexity. Additional details are available in the Supplementary Section and summarised in Fig. 2.

We used the following models to predict mortality:

1. Logistic regression model with all the original input features;
2. An autoencoder with the bio-social features as input. We then use the reduced dimensions from the autoencoder as input features to a random forest model (Fig. 2).

### Class-contrastive reasoning

We explain our models using class-contrastive reasoning and class-contrastive heatmaps. The technique works as follows. The model is trained on the training set. For each patient in the test set, we independently mutate (change from 0 to 1, or 1 to 0) each categorical feature. For each patient in the test set, we use the trained model to compute the change in the predicted probability of death.

We repeat this procedure independently for each feature and each patient in the test set. We do not retrain the model when we mutate the features. The predictions are made using the trained machine learning model on the test set.

We visualize the amount of change in the model predicted probability of mortality, achieved by setting a particular feature to 1 versus 0, using a class-contrastive heatmap. The rows represent patients and columns represent the feature that has been changed from 0 to 1. The heatmaps also show a hierarchical clustering dendrogram, which is performed using an Euclidean distance metric and complete linkage [17].

In another variant, we also simultaneously change all pairs of features in the test set from 0 and 0 to 1 and 1. As before, for each patient in the test set, we use the trained model to compute the change in the predicted probability of death. In this case, the class-contrastive heatmap shows the amount of change in the predicted probability of mortality, achieved by setting a particular combination of features to 1 versus 0. The rows represent patients and columns represent the combination of features that are changed simultaneously.

The class-contrastive heatmap shows patient-specific predictions. Predictions for individual patients are made in the following way: the trained model makes a prediction for the probability of death based on the modified features as input. This process is repeated for each patient and each feature (or feature combination).

### Survival analysis and standardized mortality ratios

For survival analysis, we use the entry date (exposure) as the date of referral. In cases where there were multiple referrals for a patient, we considered the earliest date. If this calculated date was earlier than the start date of our mental health clinical database (called RiO), we set it to the start date of RiO (1st December 2012). The event was death. The date of death was derived from the National Health Service (NHS) Spine.

We used a Cox proportional hazards model for patients with schizophrenia, using age (feature scaled) and the bio-social features (as outlined before) as input features.

We calculated age-standardized mortality ratios (SMR) to standardise and control for age and population structure. For calculating SMRs, we defined five-year age groups (0-4, 5-9, …, 85-90, and *>* 90 years). Population mortality data was used from the Office for National Statistics (ONS) [12]. We calculated SMRs for patients with a coded diagnosis of schizophrenia. SMRs were calculated using the indirect method of standardisation [18]. The denominator is the expected number of deaths in the study population and the numerator is the number of observed deaths in the study population. 95% confidence intervals were calculated as described in [18]. Additional details are available in the Supplementary Section.

### Logistic regression models

We used a logistic regression model to predict mortality in patients with schizophrenia. Age (feature scaled) and the bio-social factors were used as input. The model, in R notation, was as follows:

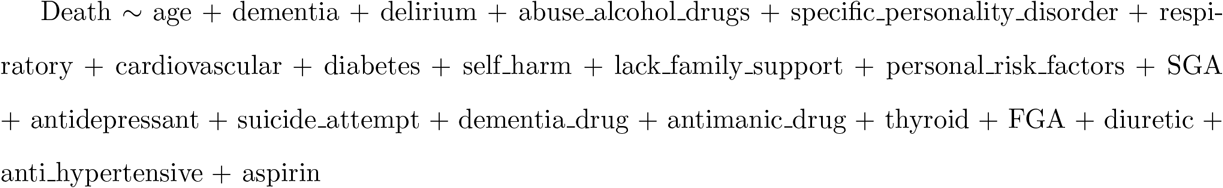

This same model was also fit using an *L*_1_ regularized logistic regression model (details are available in the Supplementary Section, Subsection Sensitivity analysis).

We also fit a logistic regression model with main effects and an interaction term between dementia in Alzheimer’s disease and cardiovascular disease. The model, in R notation, was as follows:

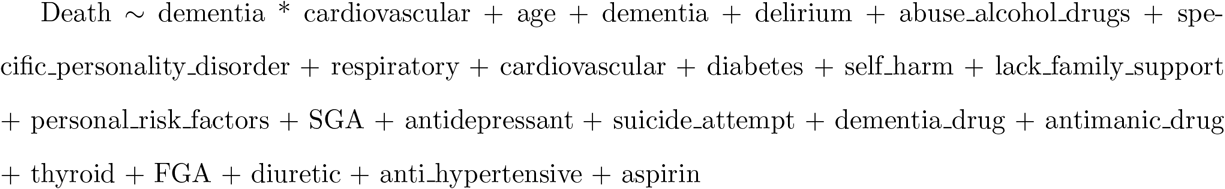

### Software

All software was written in the R [19] and Python programming languages. Visualizations were performed using the *ggplot* package in R [20]. Generalized linear model (GLM) regression was performed using the *glm* function in R [21] [15]. Hierarchical clustering and visualization were performed using heatmaps in the *pheatmap* package [22]. Survival analysis was conducted using the *survminer* package in R [23]. *L*_1_ regularized logistic regression was performed using the *glmnet* package [24].

### Data availability

This study reports on human clinical data which cannot be published directly due to reasonable privacy concerns, as per NHS research ethics approvals and information governance rules.

### Code availability

The code used in this study is available from the corresponding author upon reasonable request.

## Results

### Summary of results

We used a range of statistical and ML techniques to predict mortality in patients with schizophrenia. Class-contrastive reasoning and class-contrastive heatmaps were used to generate human-oriented explanations of statistical and ML model predictions.

Abuse of alcohol and drugs, and a diagnosis of delirium were risk factors for mortality (across all our techniques). Use of antidepressants was associated with lower risk of death via all our techniques.

The machine learning models emphasized combinations of features (like Alzheimer’s disease) with other co-morbidities. This highlights the role of co-morbidities in determining mortality in patients with SMI and the need to manage them.

### Survival analysis and standardized mortality ratios

Our explainable machine learning techniques complement classical statistical analysis like survival models and standardised mortality ratios. In this section, we outline these approaches.

For survival analysis, we use age (feature scaled) and the bio-social features (as outlined in Subsection Data input to statistical algorithms) as input features for patients with schizophrenia. For patients with schizophrenia, we show the hazard ratios associated with each feature in Fig. 3 using a Cox proportional hazards model. Use of second-generation antipsychotics (SGA) and antidepressants was associated with reduced risk of death in patients with schizophrenia. Alcohol/substance abuse was associated with an elevated risk of death consistent with a previous study [25]. A diagnosis of delirium was similarly associated with increased mortality.

**Figure 3.**
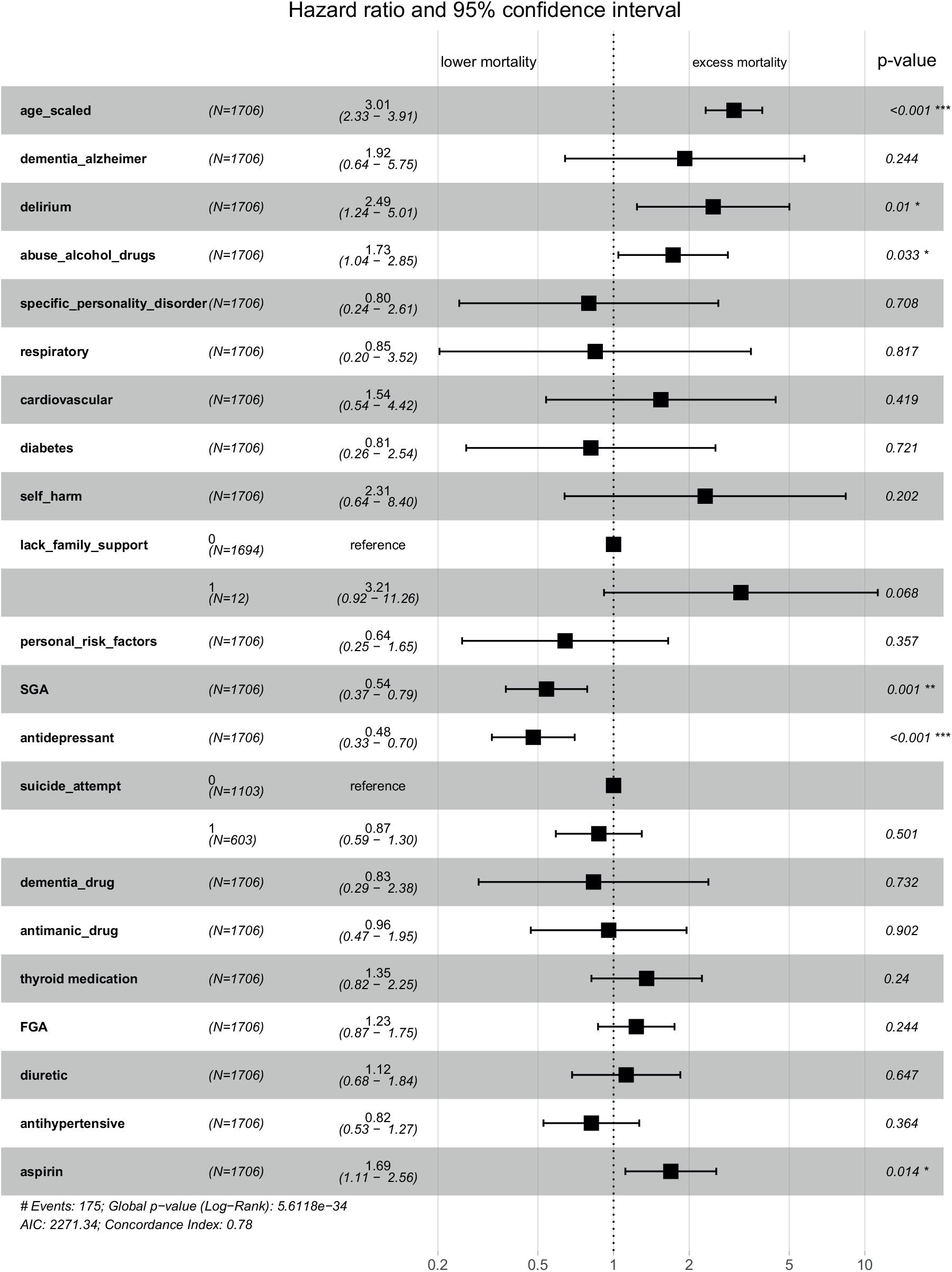
Survival analysis for patients with schizophrenia using a Cox proportional hazards model. We show the hazard ratios associated with each feature and the confidence intervals. Use of second-generation antipsychotics (SGA) and antidepressants was associated with reduced risk of death in patients with schizophrenia. Alcohol/substance abuse and a diagnosis of delirium was associated with increased mortality.

The standardized mortality ratio (SMR) for patients with schizophrenia was 7.4 (95% confidence interval: [5.5, 9.2]). This is consistent with SMRs reported in the UK [25]. Additional details are available in the Supplementary Section (Subsection Calculation of standardized mortality ratios).

### Logistic regression models

We used a logistic regression model to predict mortality in patients with schizophrenia. We show the odds ratios and their confidence intervals in Fig. 4. Age, diagnosis of delirium and alcohol/substance abuse were associated with a high risk of death. Use of second-generation antipsychotics and antidepressants were associated with a reduced risk of death.

**Figure 4.**
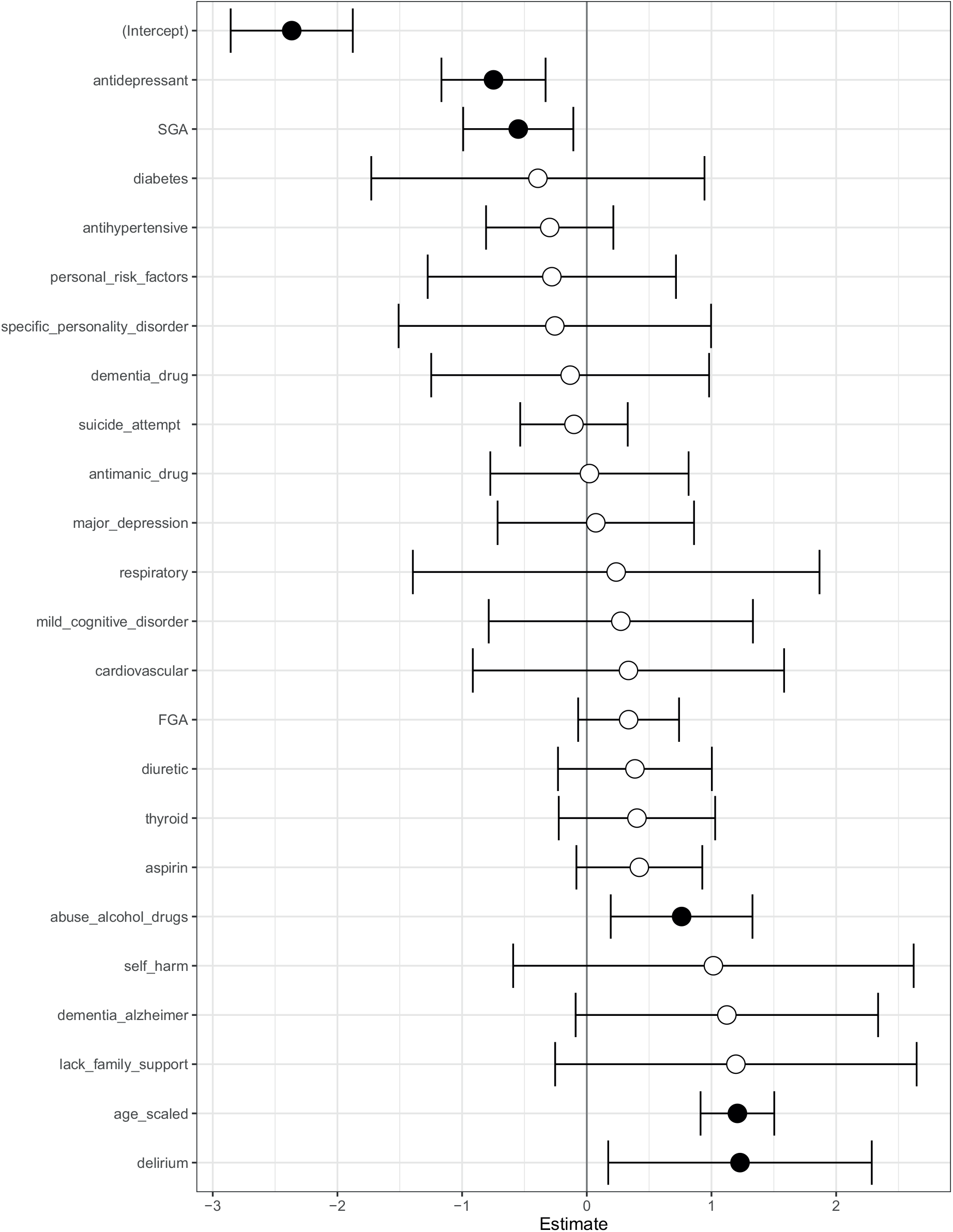
Logistic regression model to predict mortality in patients with schizophrenia. Log odds ratio of features in a logistic regression model to predict mortality in patients with schizophrenia. Shown are confidence intervals and statistical significance (filled dark circles: p-value *<* 0.05, open circles: not significant). Age, alcohol/substance abuse and a diagnosis of delirium were associated with a high risk of death. Use of second-generation antipsychotics (SGA) and antidepressants were associated with reduced risk of death.

### Class contrastive heatmaps and counter-factual statements for logistic regression

The class-contrastive explanatory technique is applicable to machine learning models and statistical models such as logistic regression. We first demonstrate our approach by using class-contrastive reasoning on the logistic regression model for predicting mortality. We show the amount of change (predicted by the trained logistic regression model on the test set) in the probability of death by changing one particular feature from 0 to 1 (in the test set). We visualize this using a heatmap (Fig. 5) where rows represent patients and columns represent features in the test set that have been changed. Predictions are made using the trained logistic regression model on the test set.

**Figure 5.**
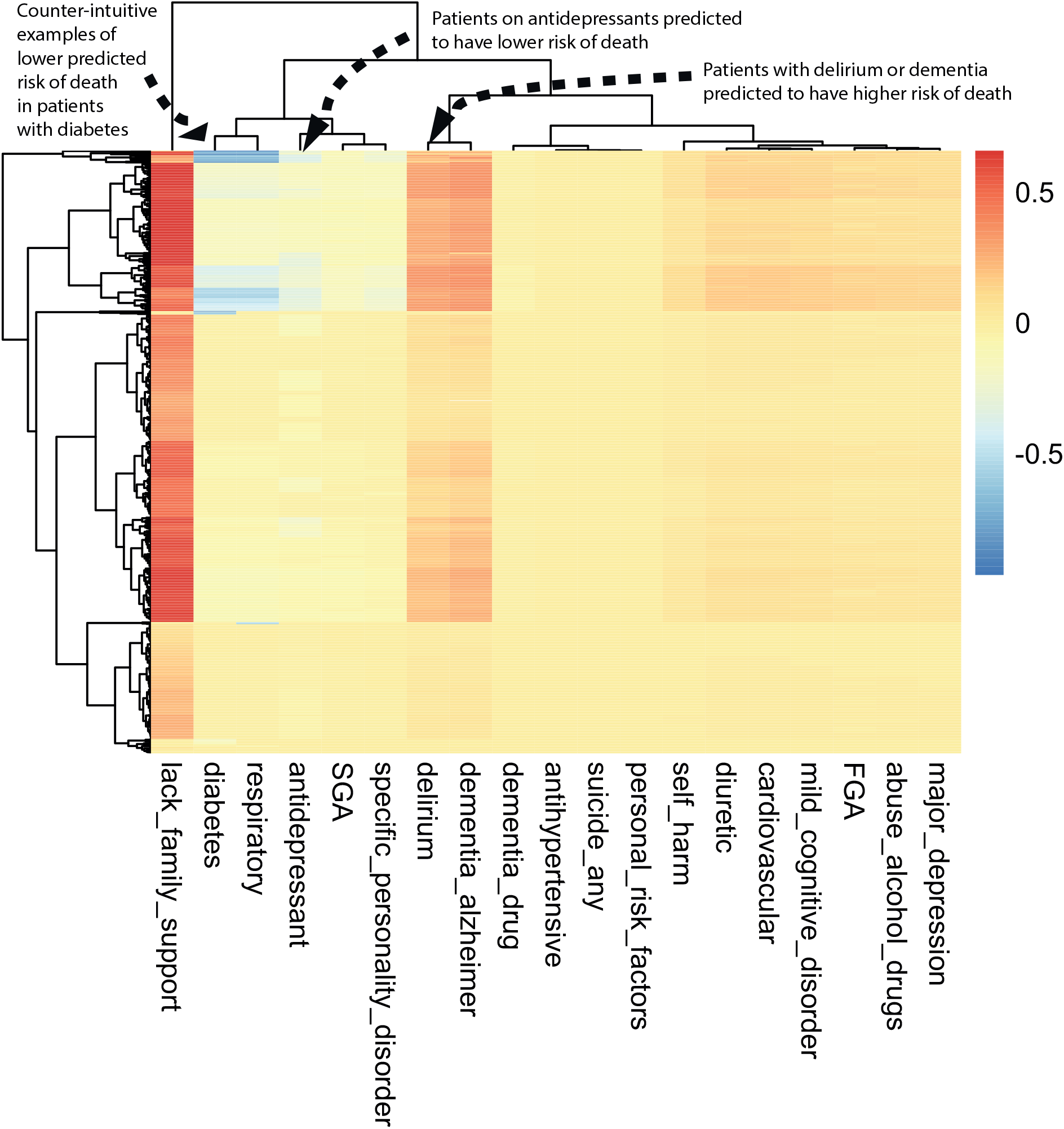
Class-contrastive heatmap for the logistic regression model. Visualization of the amount of change predicted in the probability of death by setting a particular feature to 1 versus 0 (using a logistic regression model). Rows represent patients and columns represent features. Predictions are made on the test set using the trained logistic regression model. The arrows indicate groups of patients with low predicted risk of death (shown in blue on the heatmap) and high predicted risk of death (shown in red). The arrow on the top left indicates a group of patients having a counter-intuitive characteristic of having diabetes and still have low predicted risk of death. The second arrow on the top left indicates another group of patients on antidepressants. These patients are predicted (using the logistic regression model) to have a lower risk of death. There is a third group of patients with delirium or dementia in Alzheimer’s disease (shown with an arrow) who are predicted to have a higher risk of death. The heatmap also shows a hierarchical clustering dendrogram which is performed using an Euclidean distance metric and complete linkage.

The class-contrastive heatmap shows patient-specific predictions. Predictions for individual patients are made in the following way: the trained logistic regression model makes a prediction for the probability of death based on the modified features as input. This process is repeated for each patient and each feature.

We observe that a diagnosis of delirium or dementia predisposes a group of patients towards a higher probability of predicted mortality (Fig. 5). Patients (with schizophrenia) who were taking antidepressants were less likely to die during the period observed (Fig. 5). The class-contrastive and counterfactual analysis suggests that antidepressants may be associated with lower mortality in a group of patients (Fig. 5).

The heatmap also highlights counter-intuitive predictions. For example, the heatmap suggests that there is a small sub-group of patients (Fig. 5: top left hand corner indicated with an arrow) who have diabetes and have a lower risk of death. The use of a probability scale illustrates that the effect of each predictor varies in terms of its effect on probability (according to the baseline probability determined by other variables); of course, in log odds terms, changes in a given predictor will have a constant effect across all subjects.

We note that the counter-intuitive observations we observe in the class-contrastive heatmaps (on the test set) may also be as a result of imbalances in the training set. For example, a particular binary feature may be 0 for 100 patients and 1 for 10 patients.

In order to address this, we can add synthetic training data with these imbalances and visualize the class-contrastive predictions on the test set. We can artificially introduce an imbalance (for example, add more zeros than ones to a binary feature) in the test set and training set, and then observe the class contrastive heatmaps.

We note that age is a predictor in all models that we use. However the class-contrastive heatmaps do not include age. This is because the class-contrastive analysis changes features one at a time (or pairwise), and this can be achieved only for binary categorical features. Hence, the class-contrastive heatmaps show the effect of changing predictors on the model predicted probability of mortality, over and above the contribution of age.

### Class-contrastive analysis for machine learning models

We used artificial neural networks to predict mortality in patients with schizophrenia. We performed class-contrastive analysis for this machine learning model (Fig. 2) to make it explainable.

We first show a heatmap for a simple version of class-contrastive reasoning where we mutate only one feature at a time on the test set (Fig. 6). We show the amount of change (predicted by the trained model on the test set) in the probability of death by setting one particular feature to 1 versus 0. We visualize this using a heatmap as before, where rows represent patients and columns represent features. We note that even though we cluster the features, our aim is not to demonstrate any similarity between them.

**Figure 6.**
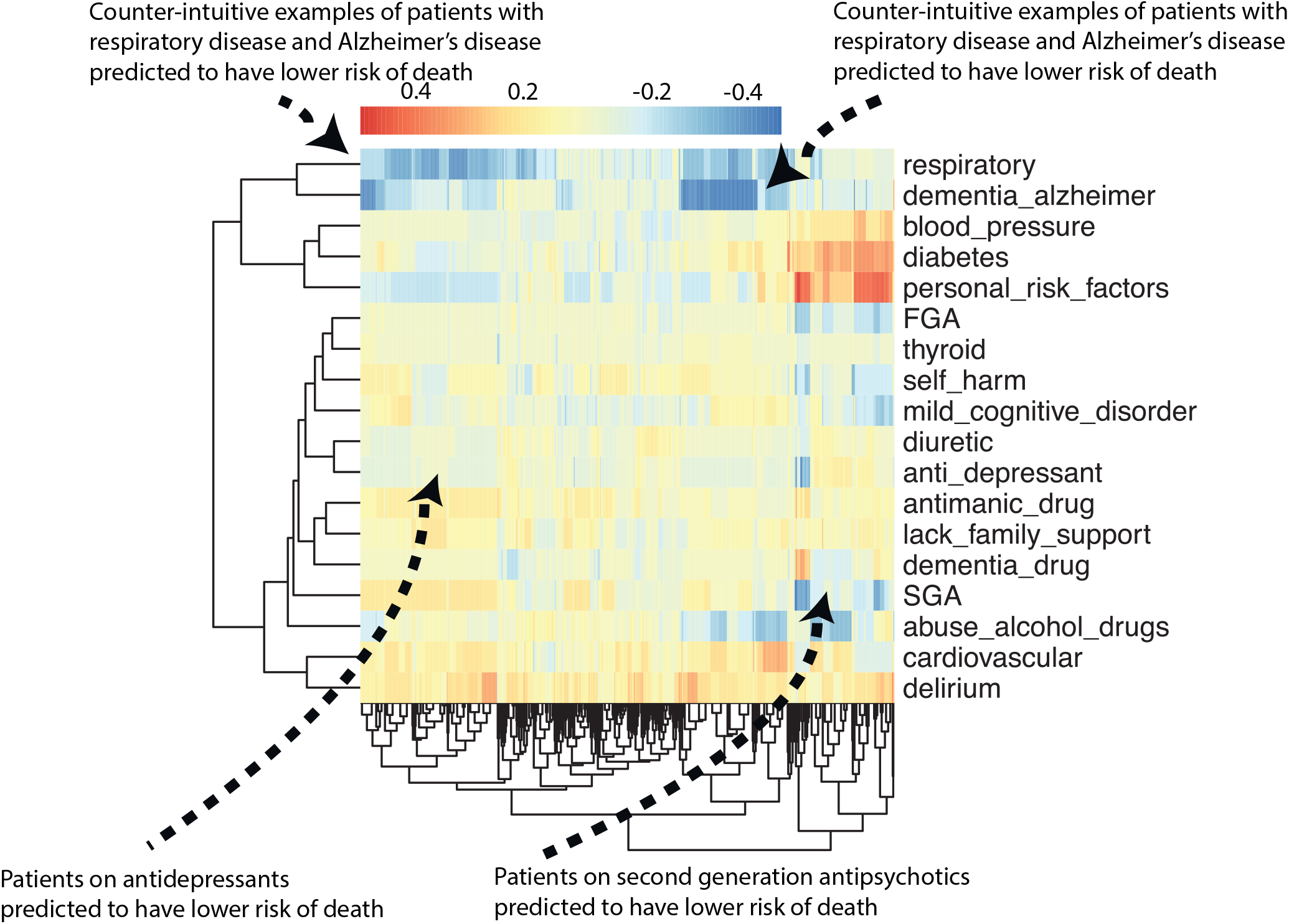
Class contrastive heatmap for the deep learning model. Visualization of the amount of change predicted in the probability of death by setting a particular feature to 1 versus 0. Predictions are made on the test set using a random forest model built on top of the autoencoder. Columns represent patients and rows represent features. The arrows at the top indicate counter-intuitive examples. If these patients had a respiratory diseases or Alzheimer’s disease, the model predicts low risk of death. The arrows at the bottom indicate a group of patients on antidepressants and SGA who are predicted to have low risk of death. The heatmap also shows a hierarchical clustering dendrogram which is performed using an Euclidean distance metric and complete linkage. We note that even though we cluster the features (columns) we do not aim to imply any similarity between them.

The heatmap suggests there is a subgroup of patients in whom use or prescription of medications like second-generation antipsychotics (SGA) and antidepressants is associated with a lower risk of death (Fig. 6). There is another subgroup of patients in whom personal risk factors (ICD-10 coded diagnosis; see Methods) are associated with increased risk of mortality.

The class-contrastive heatmaps also reveal counter-intuitive aspects of the data and model. Looking at the effect of individual features in isolation in Fig. 6, we observe small sub-groups of patients in whom having respiratory diseases or having Alzheimer’s disease is associated with a lower risk of death (indicated with arrows in Fig. 6).

These counter-intuitive results may be due to the fact that the class-contrastive approach is sensitive to the training data and any imbalances in features. For example, a binary feature may have mostly zeros in the training set. This can lead to a counter-intuitive result on the test set. Correlations across features may also help explain these counter-intuitive results.

We show an additional representative class-contrastive heatmap for the ML model in the Supplementary Section (Supp. Fig. 1). This ML model was run using a different split of the training and test data. This heatmap is consistent with previous results (Fig. 6), with the exception that it shows SGA are associated with an increased probability of mortality (Supp. Fig. 1, bottom left arrow). This is not consistent with previous results from the logistic regression model and survival analysis for the effect of SGA (Figs. 4 and 3).

Deep learning models combine input features to create higher-order representations using hidden layers. Features are also often correlated and there are non-linearities involved. To account for some higher-order (non-linear) correlations and to better highlight the combinations of features, we simultaneously change all possible combinations of two features from 0 to 1 (in the test set). Specifically, we set a particular combination of two features to 1 simultaneously (versus 0) in the test set. We then repeat this for all possible pairs of features in the test set. We visualize the change in model output on the test set in Fig. 7. This technique can be used to investigate the role of combinations of different features that deep learning models exploit to build higher-order representations.

**Figure 7.**
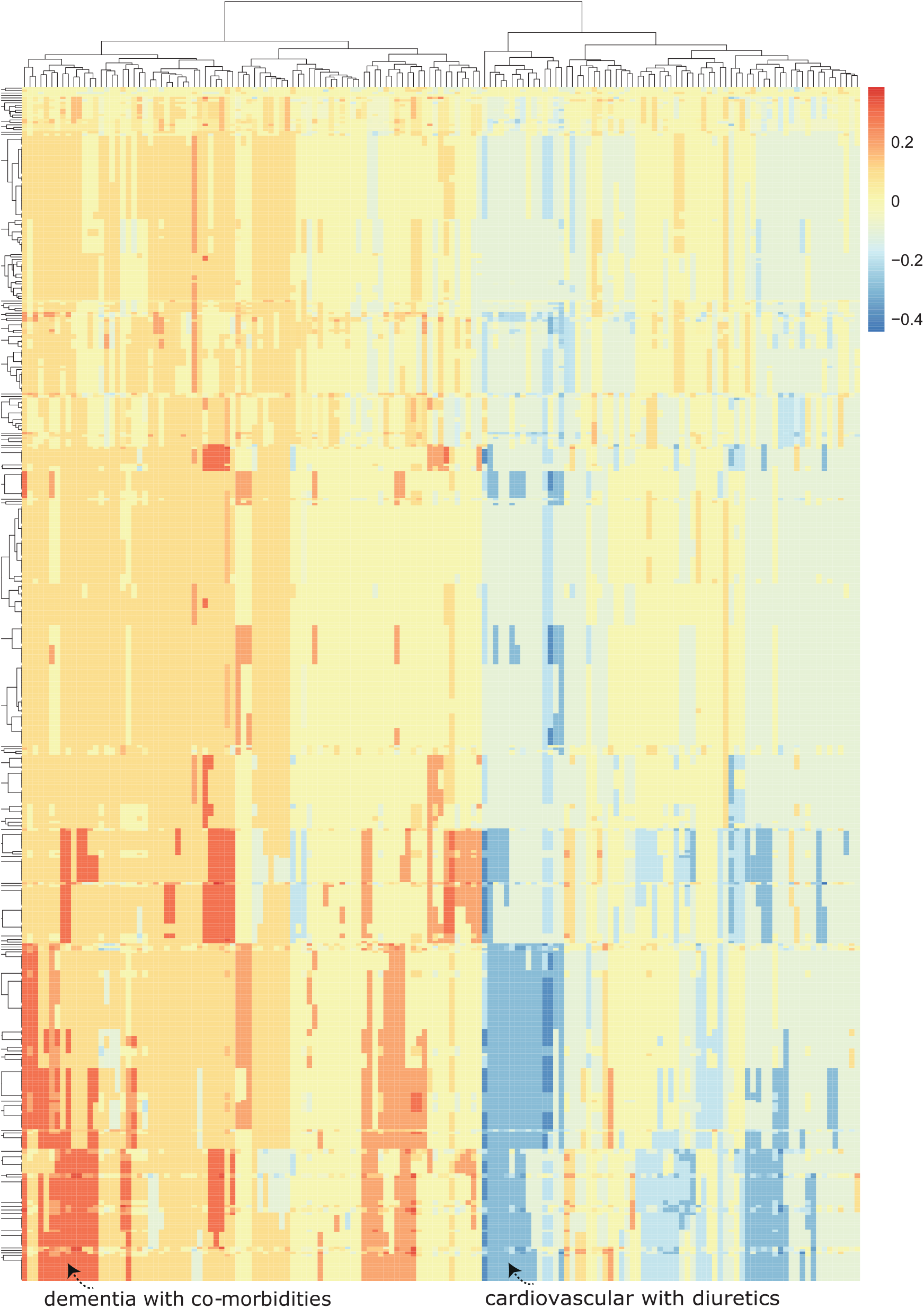
Advanced class-contrastive heatmap for the deep learning model. Class contrastive heatmap for deep learning models showing the effect of combinations of features. Visualization of the amount of change predicted in the probability of death by setting a particular combination of two features to 1 simultaneously (versus 0). Rows represent patients and columns represent feature groupings (all combinations of two features). Predictions are made on the test set using a random forest model built on top of the autoencoder. Delirium and dementia in Alzheimer’s disease seem to predispose some patients towards greater risk of mortality (shown in the lower left region of the heatmap in red). Diuretics appear to be associated with lower mortality in a group of patients with cardiovascular disease (shown in the blue region in the lower right-hand corner of the heatmap). The heatmap also shows a hierarchical clustering dendrogram which is performed using an Euclidean distance metric and complete linkage.

We found combinations of cardiovascular disease and use of diuretics. Diuretic use was associated with lower risk of mortality in a group of patients with cardiovascular disease (shown in the blue region of the heatmap in the lower right-hand corner which is the region of greatest decrease in predicted probability of death) (Fig. 7). There are also combinations of delirium and dementia in Alzheimer’s disease that predispose some patients towards greater risk of mortality (shown in the lower left region of the heatmap in red) (Fig. 7).

Other co-morbidities that are together associated with greater mortality in a sub-group of patients (Fig. 7) included: dementia in Alzheimer’s disease with an additional coded diagnosis of cardiovascular disease, and dementia in Alzheimer’s disease with a coded history of abuse of alcohol and drugs.

This highlights the role of co-morbidities in determining mortality in a sub-group of patients with SMI and the need for multiple conditions to be managed simultaneously in patients. A class-contrastive statement for one of these patients in this sub-group (Fig. 7) is: “The selected patient is at high risk of mortality because the patient has dementia in Alzheimer’s disease and has cardiovascular disease. If the patient did not have both of these characteristics, the predicted risk would be much lower.”

Our deep learning models emphasize combinations of different features. Therefore, as a very simple approximation, we also fit a more complex logistic regression model with interaction effects. We fit a logistic regression model with main effects and an interaction term between dementia in Alzheimer’s disease and cardiovascular disease (Supp. Fig. 2). The log-odds ratio for this interaction term is greater than 0 although it is not statistically significant. This may suggest that there is only a small sub-group of patients in whom dementia and cardiovascular disease co-occur and predispose towards an increased risk of death. Additional details are available in the Supplementary Section.

### Performance

We show the predictive performance of each model in this section. The models we used to predict mortality are:

1. A logistic regression model with the bio-social features as input. The area under receiver operating curve (AUC) from the logistic regression model was 0.68 (95% confidence interval [0.65, 0.70]).
2. An autoencoder with the bio-social features as input. We then used the reduced dimensions from the autoencoder as input features to a random forest model. The predicted AUC from random forests built on top of the autoencoder-reduced dimensions was 0.80 (95% confidence interval [0.78, 0.82]).

We also use other statistical learning techniques to predict mortality and these are discussed in the Supplementary Section (Section Additional analysis). We do not aim to exhaustively compare all possible statistical models but merely briefly survey and analyse some techniques. Our aim is to apply class-contrastive analysis to a machine learning model and show that in some scenarios the model predictions can be explained. We note that our aim is not to demonstrate that some machine learning models can perform better than others.

## Discussion

### Overview

Mortality among patients with severe mental illnesses (SMI) is too often premature [4] [3]. Routinely collected clinical data can help generate insights that can result in more effective treatment of these patients.

We used routinely collected clinical data in an observational study to answer questions of mortality in patients with SMI. We implemented an interpretable computational framework for integrating clinical data in mental health and interrogating it with statistical and machine learning techniques.

Our framework starts with a database that is a knowledge repository of expertise. This database was created based on consultations with clinicians and maps low-level features (for example, medications such as simvastatin) to broader categories (for example, cardiovascular medication). These features are relevant for SMI and were used to predict mortality.

Our architecture captures clinical information on physical health, mental health, personal history and social predisposing factors to create a profile for a patient. We then used a number of statistical and machine learning techniques to predict mortality using these features.

We make our predictions interpretable by using class-contrastive reasoning [6] [7]. Our approach has similarities to case-based reasoning [26] and analogy-based reasoning [27], where predictions are made based on similar patient histories cases. The approach presented here complements other techniques like Shapley explanations that are used to improve interpretability of machine learning models.

### Summary of findings

We used a range of statistical and machine learning techniques to predict mortality in patients with schizophrenia. Since machine learning models may also be difficult to explain, we make them explainable using class-contrastive reasoning and class-contrastive heatmaps.

In patients with schizophrenia, abuse of alcohol and drugs, and a diagnosis of delirium were risk factors for mortality (across all techniques). Use or prescription of antidepressants and second-generation antipsychotics (SGA) were associated with lower mortality in our logistic regression and survival models. However use or prescription of SGA was associated with an increased probability of mortality in one of our ML models.

The logistic regression model predicted that Alzheimer’s disease is a risk factor for mortality. The deep learning model emphasized Alzheimer’s disease in combination with other co-morbidities. This highlights the role of co-morbidities in determining mortality in patients with SMI and the need to manage them.

### Implications of findings

The class-contrastive and survival analysis suggest that antidepressants are associated with lower mortality in a group of patients with schizophrenia. Alcohol/substance misuse was consistently associated with elevated mortality, suggesting the requirement to address the needs of so-called “dual diagnosis” patients (with SMI and comorbid substance misuse) as part of a strategy to improve life expectancy in patients with SMI.

The association between delirium and excess mortality is notable but not unexpected [2] [3] [4]. A weakness of the current family of models is their lack of temporal structure (for example, consideration of the time between delirium and death) but this finding serves to emphasize that delirium should not be taken lightly.

The association of antidepressant use with reduced mortality was unexpected but consistent across analytical methods. Our data do not support a mechanistic interpretation (for example, mode of death is not recorded in these structured clinical records) but this question would bear further investigation.

Illicit substance abuse and lack of family involvement was associated with increased risk of mortality [25]. Alcohol/substance abuse was also pointed out as a critical factor in our class-contrastive reasoning analysis and survival analysis. Provisioning of family support and involving family members and carers could be part of health management plans [28].

We hope that some of these bio-social factors can be targeted therapeutically by either patient-level interventions (like provisioning of family support [28]) or service-level improvements [29].

Overall, we observed that abuse of alcohol and drugs and a diagnosis of delirium are risk factors for mortality (in both logistic regression models and survival models). Use of SGA and antidepressants were associated with lower mortality from both our logistic regression models and survival models. This may be important given that some clinicians may hesitate to prescribe given what is known about short-to medium-term side effects of these drugs that include adverse impact on cardiovascular risk profiles. While our findings on this and other points is not conclusive evidence of causality, it is in accord with observational clinical data at the national level [30].

The machine learning model emphasized (for example) Alzheimer’s disease along with other comorbidities (Fig. 7). This highlights the role of co-morbidities in determining mortality in patients with SMI and points to the need for multiple conditions to be treated simultaneously in patients. This also suggests that a pragmatic trial of robust management of co-morbidities may be justified.

### Interpretable algorithms for personalized clinical decision making

Interpretability is a major design consideration of machine learning algorithms applied in healthcare. We made our predictions interpretable by using class-contrastive reasoning and counterfactual statements [6]. This approach has the capability to make some black-box models explainable, which might be very useful for clinical decision support systems. We demonstrate the approach here using logistic regression and artificial neural networks. These techniques could ultimately be used to build a conversational AI that could explain its predictions to a clinician.

Our work can also be used to make clinical decision support systems. This may lead to automated alerts in electronic healthcare record systems, after thorough validation in follow-up studies.

### Limitations and future work

Our study is observational in nature and we do not imply causation. Our data is a naturalistic sample from a clinic and should not be used to alter clinical practice. Our aim is to raise hypotheses that will need to be tested in randomized controlled trials.

There may also be other unknown confounds and hence causal conclusions cannot be drawn from an observational study. For example, a drug that is associated with better outcomes may be preferred by clinical teams, and a drug that is associated with poorer outcomes may still be prescribed for severely ill patients because it is perceived to be effective.

There may also be under-coding of schizophrenia diagnoses. The data was from a secondary care mental health service provider and may miss important risk factors coded in primary care.

Psychiatric diagnoses are challenging and there can be potential issues related to the reliability of diagnostic categories in SMI. Sampling bias is another issue in real world electronic healthcare record data. For example, it is possible that only the most severely ill patients seek clinical help and get referred to secondary care. Hence the data may reflect a category of patients who are more severely ill.

Our data also lacks temporal structure which is likely to be important in determining progression of disease. The current work relates observable features to the risk of death within the observation period. Clearly this is not as satisfactory as a model that predicts a time-based risk. It would be expected that the temporal risk conferred by different features would vary – for example, diabetes increases cardiovascular risk over decades, whereas delirium is often associated with critical illness and may be associated with an elevated mortality risk that is very immediate or proximal. A comprehensive model might involve autodiscovery of those temporal risk factors, at the price of a considerable increase in model complexity. This will require building more complex recurrent neural network models like long short-term memory models (LSTM), which will require even more data.

Important readouts like statistical significance cannot be judged from the class-contrastive heatmaps. For example, lack of family support appeared to be associated with higher mortality in the class-contrastive heatmap for the logistic regression model (Fig. 5). However this association was not statistically significant, even though the odds ratio for lack of family support is greater than 1 in a logistic regression model (Fig. 4).

We combined several medications into the category of second-generation antipsychotics, which itself consists of a heterogenous group of medications [30], and make other simplifications in our treatment of medications.

Because of heterogeneity in training data and correlations across features, reproducibility of heatmaps is a limitation. We show an additional representative example in Supp. Fig. 1. There are a few differences between these heatmaps (Supp. Fig. 1 and Fig. 6). This heatmap is consistent with previous results (Fig. 6), with the exception that it shows SGA are associated with an increased probability of mortality (Supp. Fig. 1, bottom left arrow). This is not consistent with previous results from the logistic regression model and survival analysis for the effect of SGA (Figs. 4 and 3). Reconciling these results will require additional analysis and validation in an independent cohort with more patients.

Our results suggest that the class-contrastive approach is sensitive to the training data and any imbalances in features. For example, a particular binary feature may be 0 for 100 patients and 1 for 10 patients. One way to determine this sensitivity is to artificially introduce more zeros and then observe the class contrastive heatmaps.

It is possible that the counter-intuitive observations we see in the class-contrastive heatmaps (on the test set) are likely as a result of such imbalances in the training set. Because of this reproducibility of heatmaps is a limitation of our approach.

Our approach can be helpful when explicit causal structure is modelled, and when there are binary (categorical) features and a few features which can be modified at a time. We account for correlations between features by modifying all pairs of features at a time and then observing the effect on model predictions (Fig. 7). However, this approach can become computationally challenging for higher combinations of features (all triples, quadruples, all possible combinations), or as the number of features increase.

### Concluding remarks

Our framework combines bio-social factors relevant for SMI with statistical learning, and makes them interpretable using class-contrastive techniques. Our work suggests that medications like antidepressants were associated with a reduced risk of death in a group of patients with schizophrenia. Abuse of alcohol and drugs, and a diagnosis of delirium were risk factors for death.

Our machine learning models highlight the role of co-morbidities in determining mortality in patients with SMI and the need to manage them. We hope that some of these bio-social factors can be targeted therapeutically by either patient-level or service-level interventions.

We complement explainable machine learning techniques with classical statistical analysis like logistic regression, survival models and standardised mortality ratios. This may be a prudent and pragmatic approach for building explainable models in healthcare. We admit that the distinction between ML models and classical statistical models (like logistic regression) is artificial. Models lie on a continuum and a pragmatic approach towards explainable AI would combine and contrast all of these techniques.

The approach of combining explainable techniques and clinical knowledge with machine learning approaches may be more broadly applicable when data scientists need to work closely with domain experts (clinicians and patients).

Our approach combines clinical knowledge, health data, and statistical learning, to make predictions interpretable to clinicians using class-contrastive reasoning. We view our work as a step towards interpretable AI and personalized medicine for patients with SMI and potentially other diseases.

## Acknowledgements

We thank Jenny Nelder and Jonathan Lewis for all their support during this project and Irene Egli for inspiring us to think about patients with schizophrenia. This work is dedicated to the memory of Patrick Winston.

## Ethics

The CPFT Research Database operates under UK NHS Research Ethics approvals (REC references 12/EE/0407, 17/EE/0442; IRAS project ID 237953).

## Author contributions

SB, PL, PBJ, and RNC designed the study. SB and RNC verified the underlying data. SB conducted the analyses and wrote the original draft of the manuscript. All authors edited the manuscript and gave final approval for publication.

## Supplementary Information

### Supplementary Methods

#### Machine learning methods

We used an artificial neural network, called an autoencoder, to integrate data from different sources and predict mortality. The input features are age (normalised), gender, diagnosis categories, lifestyle risk factors, social factors and medication categories. We use the same set of features for all algorithms.

Categorical features (such as medication categories) are encoded using a one-hot representation. This involves taking a vector that is as long as the number of unique values of the feature. Each position on this vector corresponds to a unique value that the categorical feature can take. Whenever a categorical feature (say, did a patient take cardiovascular medication) takes on a particular value (say True), we place a 1 (“hot”) corresponding to that position on the vector and 0 everywhere else.

We show the architecture of the autoencoder in Fig. 2. The autoencoder is an artificial neural network with an input layer, hidden layer and an output layer. The input layer takes in the bio-social features. The output layer is used to reconstruct the input. The hidden layer of the autoencoder is used for dimensionality reduction.

The autoencoder has one hidden layer of 10 neurons. We use the hidden layer as input to a random forest model to predict mortality. A similar architecture was applied previously to electronic healthcare record data [31]. The choice of an autoencoder allows reduction of the feature space.

An artificial neural network has an input layer, hidden layer(s) and output layer. An activation function is used to project the input data (*X*) into another feature space using weights (*W*).

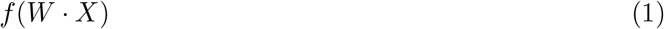

The weights *W* are determined using a technique called backpropagation [32].

We used a ReLU (Rectified Linear Unit) activation function for the hidden layer. The form of the ReLU function is shown below:

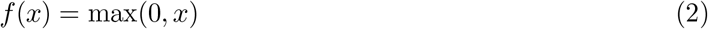

We used a sigmoid activation function for the final layer:

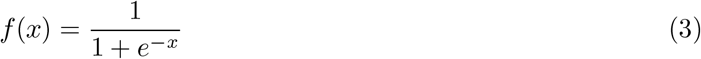

The output of the sigmoid function is positive even for negative input.

We also experimented with a hyperbolic tangent (tanh) function shown below:

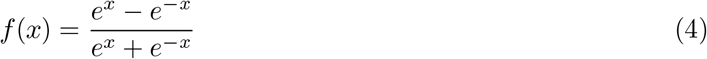

However the cross-validation results (see discussion later) were inferior to that of the ReLU activation function.

An artificial feed-forward neural network optimizes a loss function of the form:

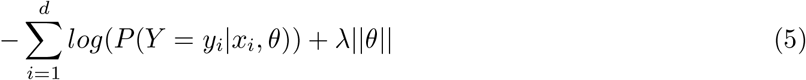

This is the negative log-likelihood. There are *d* data points. The *i* th data point has a label denoted by *y*_*i*_ and input feature vector represented by *x*_*i*_. The weights of the artificial neural network are represented by a vector *θ. λ* is a regularization parameter to prevent overfitting and reduce model complexity. *λ* is usually determined by cross-validation. Shown here is the *L*_1_ norm of the parameter vector (*θ*). The weights of the artificial neural network are determined using a technique called backpropagation [32].

The autoencoder used a cross-entropy loss function, which is a measure of discrepancy between the input layer and reconstructed hidden layer. The cross-entropy loss function used for the autoencoder had the following form:

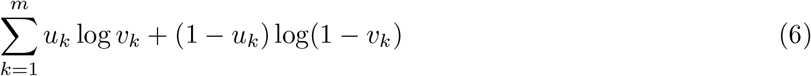

where there are *m* features in the input layer. *u* represents the input layer and *v* represents the hidden layer. The layers are computed by applying the appropriate activation functions (Equations 1, 2 and 3).

The final cost function is given below:

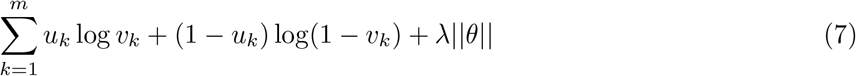

where the vector *θ* represents all the weights of the artificial neural network. There are *m* features in the input layer. *u* represents the input layer and *v* represents the hidden layer. We added an *L*_1_ penalty term on the weights to perform regularization and prevent overfitting. This is denoted by the term *λ*||*θ*||. *λ* is a regularization parameter which we determined by 10-fold cross-validation.

We performed a 50%-25%-25% training-validation-test split of the data. We used the *keras* package [33] with the *Tensorflow* backend [34].

The artificial neural network was trained on the training data for a number of epochs. In one epoch the network is trained on the training dataset. The model fit is then refined over subsequent epochs. Our neural network was trained for 1000 epochs, which was assessed as being sufficient to reach convergence. We used the *Adadelta* method of optimization [35].

We selected all hyperparameters, including the number of neurons in a hidden layer and activation functions, based on a uniform search and 10-fold cross-validation. We split the data into training set (50%), validation set (25%) and test set (25%). We trained the model on the training set. We carried out cross-validation on the validation set. The architectural parameters and regularization parameters are then selected. This final model is then evaluated on the test set. This process of splitting the data (into training, validation and test sets), training the model and performing cross-validation is repeated 10 times.

We varied the number of neurons in the hidden layer from 2 to 20. For activation functions, we tried sigmoid, rectified linear unit (ReLU) and hyperbolic tangent (tanh). We do not use dropout regularization to keep a simple architecture and simplify the process of model selection. A hidden layer of 10 neurons and ReLU and sigmoid activation functions (for the first and second layers, respectively), were found to have the least cross-validation error.

We repeated the stochastic process of splitting the data into training and test sets and performing cross-validation 10 times. This yielded a mean AUC of 0.80 (95% confidence intervals [0.78, 0.82]).

#### Calculation of standardized mortality ratios

Standardized mortality ratios (SMR) are a method to standardise and control for age and population structure [18]. We calculated SMRs using the indirect method of standardisation [18]. The denominator is the expected number of deaths in the study population and the numerator is the number of observed deaths in the study population.

Hence the indirectly standardised SMR is the ratio of the number of deaths observed in a study population to the number expected if the age-specific rates of a standard population had applied:

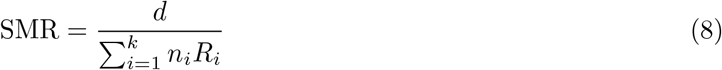

where *d* is the number of deaths in the study population. Say there are *k* age groups in the study and standard population. *n*_*i*_ is the number of people in the *i*th group of the study population and *R*_*i*_ is the crude death rate in the *i*th group of the standard population. The 95% confidence intervals are SMR ± 1.96 · SE(SMR) [18] where SE(SMR) is given by:

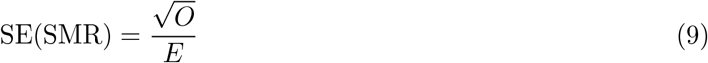

Here *O* is the observed number of deaths in the study population and *E* is the expected number of deaths in the study population.

### Sensitivity analysis

#### Additional analysis

For the ML model (random forest built on features from the autoencoder), we performed additional sensitivity analysis. We repeated the stochastic process of splitting the data into training and test sets and performing cross-validation 10 times. We then performed stability analysis of heatmaps. We generated heatmaps for each of the 10 iterations mentioned above. We show a representative heatmap in Supp. Fig. 1.

Because of heterogeneity in training data and correlations across features, reproducibility of heatmaps is a challenge. We acknowledge this limitation. We show a representative example in Supp. Fig. 1. This heatmap is consistent with previous results (Fig. 6), with the exception that it shows use or prescription of SGA is associated with an increased probability of mortality (Supp. Fig. 1, shown by arrow on bottom left corner). This is not consistent with previous results from the logistic regression model and survival analysis (Fig. 4 and Fig. 3). Reconciling these results will require additional analysis in an independent cohort with more patients.

Consistent with the previous results (Fig. 6), this new heatmap (Supp. Fig. 1) also shows the counter-intuitive result that for some patients with respiratory disease or Alzheimer’s disease, the model predicts a lower risk of death.

Our results suggest that the class-contrastive approach is sensitive to the training data and any imbalances in features (e.g. a particular binary feature may be 0 for 100 patients and 1 for 10 patients). Correlations across features may also help explain these counter-intuitive results.

We also used the following models to predict mortality: 1) a random forest model operating on the original features (95% CI of AUC: [0.71, 0.79]), 2) performing PCA on the original features and using these reduced dimensions as features to a random forest model (95% CI of AUC: [0.51, 0.76]) and logistic regression model (95% CI of AUC: [0.52, 0.77]), and 3) *L*_1_ regularized logistic regression model using the original features (95% CI of AUC: [0.72, 0.74]). We performed PCA on the original input features. The top 10 principal components were then used as input to a logistic regression model and (independently) a random forest model.

For the *L*_1_ regularized logistic regression model, we optimized the regularization hyperparameter as described before. Briefly, we split the data into training set (50%), validation set (25%) and test set (25%). We trained the model on the training set. We carried out cross-validation on the validation set. The regularization parameter for an *L*_1_ penalized logistic regression model is then selected. This final model is then evaluated on the test set. This process of splitting the data (into training, validation and test sets), training the model and performing cross-validation is repeated 10 times.

Finally, we also fit a *L*_1_ regularized logistic regression model where age was divided by 100 (instead of being scaled by subtracting the mean and dividing by the standard deviation). This model had similar predictive power compared to a model where age was standardized (95% CI of AUC [0.69, 0.73]). Hence we use the standardization method throughout for the age variable (subtracting the mean and dividing by the standard deviation).

Our aim is not to exhaustively compare all possible statistical models but merely briefly survey and analyse some techniques. Our aim is to apply class-contrastive analysis to a few machine learning models and show that in some scenarios the model predictions can be explained. We note that our aim is not to demonstrate that machine learning models can perform better than others.

Our objective is not to show that a particular ML algorithm is better but to show that ML approaches can be made interpretable in some scenarios using class-contrastive reasoning. We show a practical demonstration on a clinical dataset in a disease of public health relevance.

#### Logistic regression models with interaction effects

Our deep learning models emphasize combinations of different features. Hence, as a very simple approximation, we also fit a more logistic regression model with interaction effects and main effects.

We fit a logistic regression model with main effects and an interaction term to account for comorbidities: dementia and cardiovascular disease (Supp. Fig. 2).

The model, in R notation, was as follows:

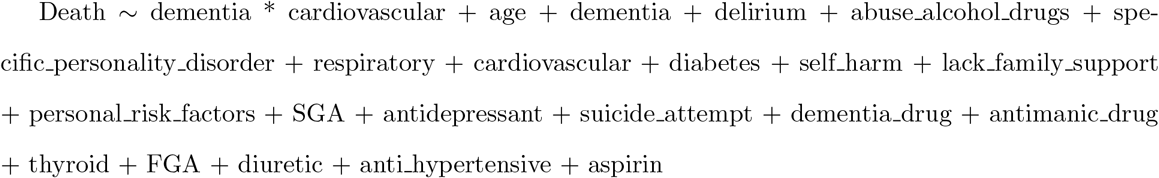

We also fit a logistic regression model where age interacts with all other features (Supp. Fig. 3). The model is:

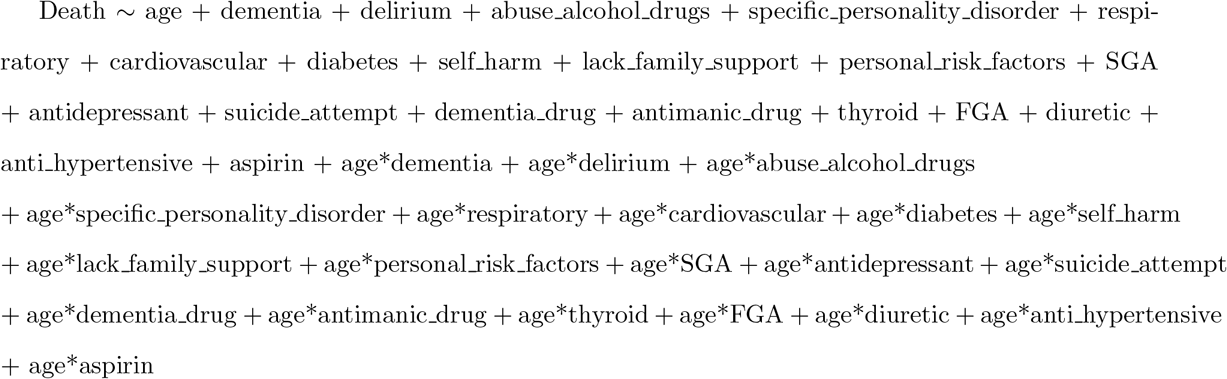

## Supplementary Figures

**Supplementary Figure 1.**
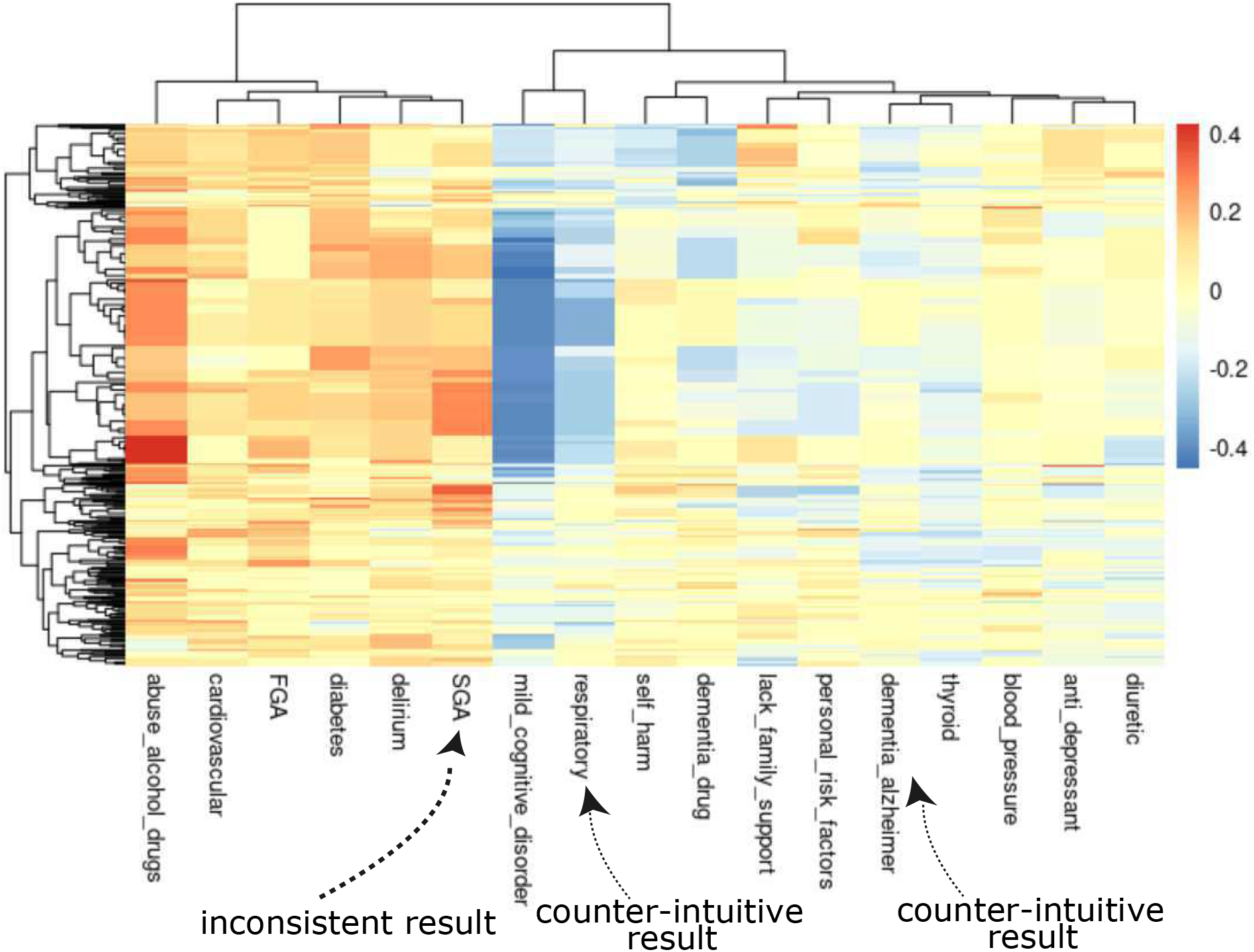
Sensitivity analysis for class contrastive heatmap for the deep learning model. Visualization of the amount of change predicted in the probability of death by setting a particular feature to 1 versus 0. Predictions are made on the test set using a random forest model built on top of the autoencoder. Columns represent patients and rows represent features. The arrows at the bottom right indicate counter-intuitive examples. If these patients had a respiratory diseases or Alzheimer’s disease, the model predicts low risk of death. The arrow at the bottom left indicates a group of patients on SGA who are predicted to have high risk of death. This is inconsistent with analysis from the logistic regression and survival models. The heatmap also shows a hierarchical clustering dendrogram which is performed using an Euclidean distance metric and complete linkage. We note that even though we cluster the features (columns) we do not aim to imply any similarity between them.

**Supplementary Figure 2.**
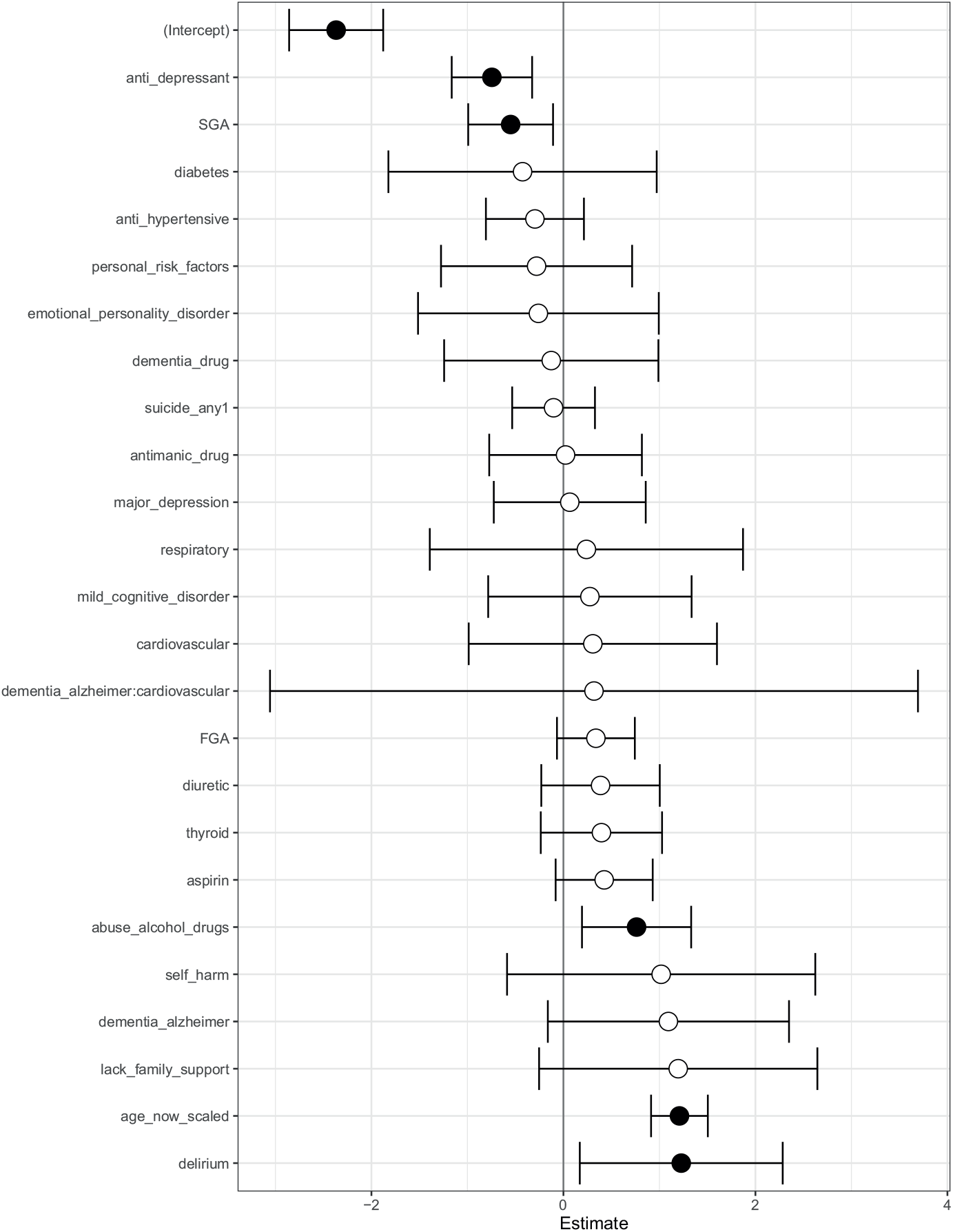
A logistic regression model with main effects and an interaction effect between dementia and cardiovascular disease. Log odds ratio for each feature from a logistic regression model for predicting mortality in patients with schizophrenia. The logistic regression model has main effects and an interaction between dementia and cardiovascular disease. Shown are confidence intervals and statistical significance (filled dark circles: p-value *<* 0.05, open circles: not significant).

**Supplementary Figure 3.**
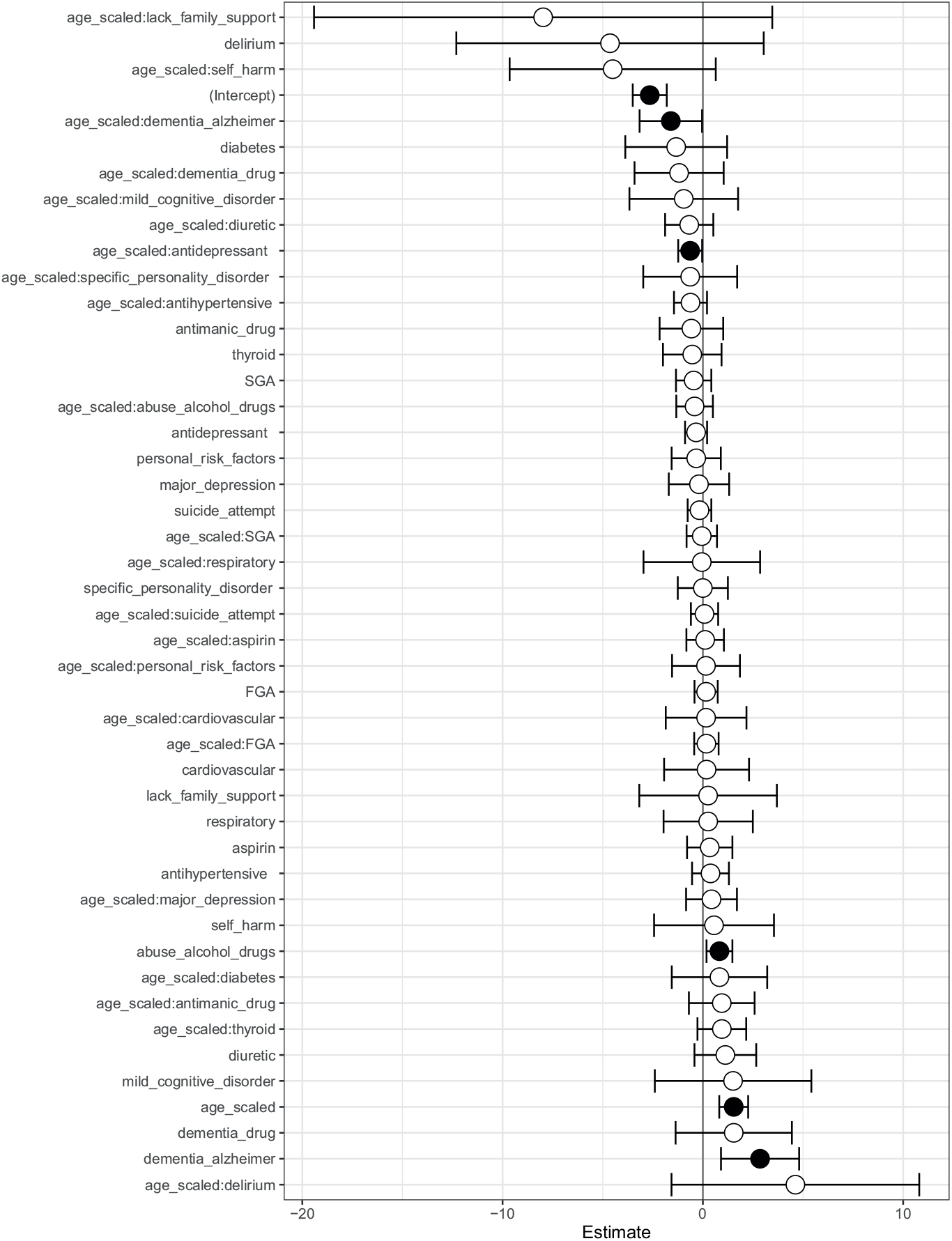
Logistic regression model with main effects and all pairwise interactions with age. Log odds ratio for each feature from a logistic regression model for predicting mortality in patients with schizophrenia. The logistic regression model has main effects and all pairwise interactions with age. Shown are confidence intervals and statistical significance (filled dark circles: p-value *<* 0.05, open circles: not significant).

